# Bridging the Gap: Socioeconomic Disparities in Coronary Heart Disease Knowledge, Awareness, and Lifestyle Practices in Bangladesh

**DOI:** 10.1101/2025.09.01.25334836

**Authors:** Nishat-E-Sharmin Trisha, Herbert F. Jelinek

## Abstract

**Background:** Coronary heart disease (CHD) is a leading cause of morbidity and mortality globally, with a rising burden in low– and middle-income countries such as Bangladesh due to demographic and lifestyle changes. Socioeconomic disparities influence exposure to CHD risk factors, access to health information, and adoption of preventive behaviors. Impact of these disparities is essential for designing targeted and inclusive interventions. The main aim of this study was to compare coronary heart disease (CHD)-related knowledge, awareness, and lifestyle practices between people of high socioeconomic status (HSES) and low socioeconomic status (LSES) in Dhaka, Bangladesh. The relationship between knowledge and awareness about CHD was also examined, as were relationships of knowledge and awareness with lifestyle practices.

**Methods:** A cross-sectional study was conducted using a validated four-section questionnaire, developed in English, translated into Bengali, and pretested for clarity and cultural appropriateness. Semistructured interviews, comprising both open and fixed-choice questions, were used to obtain information from 238 HSES and 240 LSES people. Knowledge concerning CHD was assessed in terms of epidemiology, pathophysiology, risk factors, symptoms of heart attack, prevention, and treatment. Participants were asked about their awareness concerning CHD, their risk factors, taking preventive action against CHD, prediagnosed medical conditions including CHD, family history of CHD, awareness of any people who had been diagnosed with CHD in their community, and the action that the participant would take in the event of a heart attack. Participants’ lifestyle practices were assessed concerning tobacco smoking, tobacco chewing, physical activity, dietary habits, and alcohol consumption. Data were analyzed with goodness-of-fit chi-square tests, correlations, and multiple linear regressions.

**Results:** None of the participants in either group were categorized as having a good or very good level of knowledge about CHD, and very low percentages of participants had either a good or very good level of awareness about CHD. However, HSES participants had higher levels of both knowledge and awareness about CHD than did LSES interviewees. More than two-thirds of the participants from both groups had lifestyle scores indicating good or very good lifestyle practices. Knowledge and awareness about CHD were associated with each other in both HSES and LSES groups, but neither knowledge nor awareness about CHD was related to lifestyle practices. The multiple regressions indicated that participants from the HSES who were older had greater knowledge about CHD; those from the HSES who were male had greater awareness about CHD; and that better lifestyle practices were evident in HSES and younger participants.

**Conclusions:** Although knowledge and awareness about CHD are related to socioeconomic status, neither of those variables is associated with healthy behaviors. These findings highlight the need for context-specific health education programs to address gaps in CHD knowledge and awareness among lower socioeconomic groups in Bangladesh.

## Introduction

Coronary heart disease (CHD) is a significant global public health issue, responsible for approximately 16.6% of global deaths in 2019 [1]. The burden of CHD is increasing in low-income countries such as Bangladesh due to the increasing prevalence of non-communicable diseases (NCDs) resulting from demographic transition [2][3]. Unfortunately, data related to CHD in Bangladesh are often insufficient and contain statistical errors, which hinder health promotion and preventive strategies [2]. Established behavioral risk factors associated with CHD, such as tobacco use, unhealthy diet, physical inactivity, and harmful alcohol consumption, are prevalent in Bangladesh and increase the likelihood of CHD [4][5][6][7][8].

Preventive strategies for CHD have been identified by government and non-governmental organizations [9]. However, understanding the level of CHD knowledge and awareness in the general population is crucial for successful primary and secondary prevention. Lack of knowledge and awareness about CHD can lead to increased prevalence and mortality rates [10]. While knowledge and awareness are related constructs, they can be distinguished from each other. Knowledge refers to factual information acquired from authoritative external sources, whereas awareness includes personalization and self-focus based on personal familiarity or concern [11][12].

Socioeconomic status (SES) is a critical factor that affects exposure and vulnerability to CHD risk factors. CHD is associated with social and economic inequalities [13]. People of high SES (HSES) and low SES (LSES) may have different levels of knowledge, awareness, and lifestyle practices related to CHD. Previous studies have indicated that people of both HSES and LSES are at risk of CHD in Bangladesh, but for different reasons [14][15][16][17][18]. For instance, HSES people tend to be physically inactive and have higher percentages of hypertension, diabetes, and overweight/obesity, while tobacco use is more prevalent among LSES people [19][20].

Bangladesh-specific evidence highlights both the increasing burden of cardiovascular risk and significant gaps in public knowledge and prevention. The 2018 national WHO STEPS survey found high rates of major modifiable risk factors among Bangladeshi adults, which included inadequate fruit and vegetable intake, low physical activity in some groups, tobacco use, overweight and obesity, hypertension, diabetes, and elevated lipids. Significantly, about a quarter of the adult population experiences three or more of these risk factors and is impacted by sociodemographic gradients [21] [22] [23].

Several studies based in Bangladesh show limited knowledge and awareness of CHD and CVD, especially among those with less education or income. A review from Bangladesh identified coronary artery disease as a leading cause of death across the country, highlighting both common and specific risks such as air pollution, arsenic exposure, and vitamin D deficiency. Surveys of populations and communities have since revealed limited awareness of symptoms and risk factors, along with poor preventive practices among the public and patient groups, with higher education and income associated with better knowledge and habits. Additionally, studies in slum areas highlight a high prevalence of hypertension and related risks, worsened by limited health literacy [24] [25] [26] [27] [28] [29].

Despite the importance of socioeconomic status in CHD prevention and management, little is known about the relationship between SES and knowledge, awareness, and lifestyle practices related to CHD in Bangladesh [30]. To address this gap in the research, this study aimed to compare people of HSES and LSES in Bangladesh in terms of their knowledge, awareness, and lifestyle practices related to CHD. The results of this study could help identify disparities in CHD prevention and management between HSES and LSES people and inform future interventions targeted at specific subgroups within the general population.

## Methods

### Study setting and rationale

This study was completed in Dhaka, where the residential and occupational groups allowed for recruiting HSES and LSES participants from workplaces and low-income settlements. Focusing data collection in one city minimized variations in procedures, language, and healthcare, improving comparability of knowledge, awareness, and lifestyle outcomes across SES groups. Dhaka was chosen to capture a broad SES spectrum within a single cultural and healthcare context, which facilitates logistics for face-to-face interviews, and targets urban areas where CHD prevention efforts are most effective.

### Participants

The study included residents of Dhaka, Bangladesh, aged between 18 and 60. The sample comprised 238 participants of high socioeconomic status (HSES) who were office workers in government and nongovernment organizations, and 240 participants of low socioeconomic status (LSES) residing in slum areas. Pregnant women and individuals with noticeable cognitive impairment, intellectual disability, or mental illness were excluded. Final data collection occurred during the 4 months from 01/09/2014 to 20/12/2014 and all participants provided written informed consent.

### Sample size and power

We conducted an a priori power analysis for the primary between-group comparisons (high socioeconomic status [HSES] vs low socioeconomic status [LSES]). Assuming a two-sided α=0.05, we needed at least 233 participants per group to detect a small standardized mean difference (Cohen’s d≈0.26) with 80% power (or approximately 90% power for d≈0.30). Based on this, we aimed for a minimum of 233 participants per group and enrolled 238 HSES and 240 LSES participants. This calculation was designed to ensure sufficient sensitivity for small SES-related differences in knowledge, awareness, and lifestyle practice scores, while accommodating practical field constraints in Dhaka.

### Questionnaire Development, Translation, and Validation

Content validity and expert review (Bangladesh): The initial English-language instrument consists of four sections, including demographics, CHD knowledge, CHD awareness, and lifestyle practices, and was reviewed by academic staff with subject-matter expertise and then evaluated by public health and cardiology experts in Bangladesh for content coverage, clarity, and cultural appropriateness. Revisions were made to ensure comprehensive coverage of CHD epidemiology, pathophysiology, risk factors, symptoms, prevention, and treatment.

Translation and back-translation: Following revision, the questionnaire was forward-translated into Bengali by a bilingual researcher and independently back-translated by a second bilingual professional. Discrepancies were reconciled by consensus. The Bengali version was then reviewed by the Research Review Committee of the Centre for Control of Chronic Diseases (Bangladesh) for linguistic and cultural fidelity.

Pretesting and pilot in Bangladesh: We conducted interviews and pilot testing with HSES (n=12) and LSES (n=8) adults to evaluate item comprehension, response options, and administration flow. Where required, minor wording and ordering of questions were changed to improve clarity and reduce interview burden.

Measurement reliability (internal consistency and stability): Internal consistency was evaluated for the multi-item Knowledge, Awareness, and Lifestyle scales. Short-interval test–retest reliability was measured in a subsample using intraclass correlation coefficients. Item-level performance was analyzed to verify stable distributions prior to the main analyses.

Administration of questionnaire: One of the Bengali-speaking researchers administered the final Bengali questionnaire face-to-face using standardized scripts to minimize interviewer effects and to support participants with low literacy (Supplement 1).

### Procedure

For HSES participants, eight major commercial areas within the 49 Dhaka city subdistricts were selected, and within each location, a building was chosen to invite participants. In the case of LSES participants, eight subdistricts were selected, starting from the middle of the list, and the closest slum area to the police station in each subdistrict was identified. Rooms within the slum areas were chosen as the physical sites, and participants were selected based on proximity and eligibility. Informed consent was obtained from participants before conducting face-to-face structured interviews, which were audio-recorded and later transcribed for analysis.

### Knowledge, Awareness, and Lifestyle Practice Scores and Categorization

Participants’ knowledge scores were calculated based on the number of correct answers across the CHD-knowledge questions. Awareness scores were determined similarly for the awareness-related questions. Lifestyle practice scores were obtained by assigning scores to different domains and summing them. The maximum scores were divided into five categories: very poor, poor, moderate, good, and very good. Each participant was placed in one of these categories based on their scores.

### Analyses

Data was analyzed using SPSS (Version 20, IBM) and Excel. Descriptive statistics were used to summarize the data. Chi-square tests were performed for categorical variables, and Bonferroni adjustments were applied for multiple comparisons. Spearman’s rank-order correlation coefficients were used to identify associations between variables. Multiple linear regression models were built to examine the relationships between variables while controlling for age and gender. The models were ranked using the Akaike information criterion (AIC), and multicollinearity was checked using the variance inflation factor (VIF).

Goodness-of-fit and significance measures were assessed before final model selection. A significance level of p < .05 was considered statistically significant, with adjusted thresholds for specific analyses. Scale reliability was determined with Cronbach’s α (internal consistency) and, where available, test–retest ICCs (two-way mixed, absolute agreement), reporting 95% CIs; assumptions and item distributions were examined before inferential tests

## Results

Demographic characteristics of the participants were used to establish two distinct socioeconomic status (SES) groups for the study. The updated version of the Kuppuswamy Socioeconomic Scale [31], initially developed in India for a population with a similar socioeconomic background to Bangladesh, was utilized to categorize the participants. This scale consists of five SES categories.

To ensure the success of the procedure in obtaining the two distinct SES groups, the researchers relied on the demographic information provided by the participants. All participants identified as likely to belong to the high socioeconomic status (HSES) group were placed in the highest category (Category I) of the updated scale. On the other hand, participants identified as likely to belong to the low socioeconomic status (LSES) group were placed in the two lowest categories (Categories IV and V).

Among the HSES participants, the majority were higher officials in insurance companies (37%) or bankers (35.3%). Other occupations represented in this group included industrialists (10.5%), senior officials in business organizations (10.1%), and higher officials in Bangladesh customs (5.0%).

In contrast, the largest percentage of LSES participants were garment workers (37.5%), followed by factory workers (16.7%), housemaids (12.5%), trade workers (8.9%), and rickshaw operators (6.2%).

The age of the HSES participants (M = 40.27, SD = 9.5 years) was found to be significantly higher than that of the LSES participants (M = 29.53, SD = 10.3 years). This difference was statistically significant, as indicated by the t-test (t(476) = 11.90, p < .001, d = 1.08), suggesting an age disparity between the two SES groups.

Furthermore, there was a notable gender disparity within the HSES group, with a significantly higher proportion of male participants (71.4% males vs. 28.6% females). This gender imbalance reflected the characteristics of the workplaces from which the HSES data had been collected. In contrast, the proportion of males versus females in the LSES group was less imbalanced, with 54.6% males and 45.4% females.

Additionally, educational attainment differed significantly between males and females in the LSES group. Among females, a higher percentage (30.3%) had never attended school compared to males (10.7%). This difference was determined to be statistically significant, as indicated by the chi-square test (χ2 (1, N = 240) = 14.44, p < .001). Table 1 presents the occupation distribution, percentage, average age with standard deviation, and gender composition (specifically, the percentage of participants who never attended school) for each SES group (HSES and LSES).

**Table 1.**
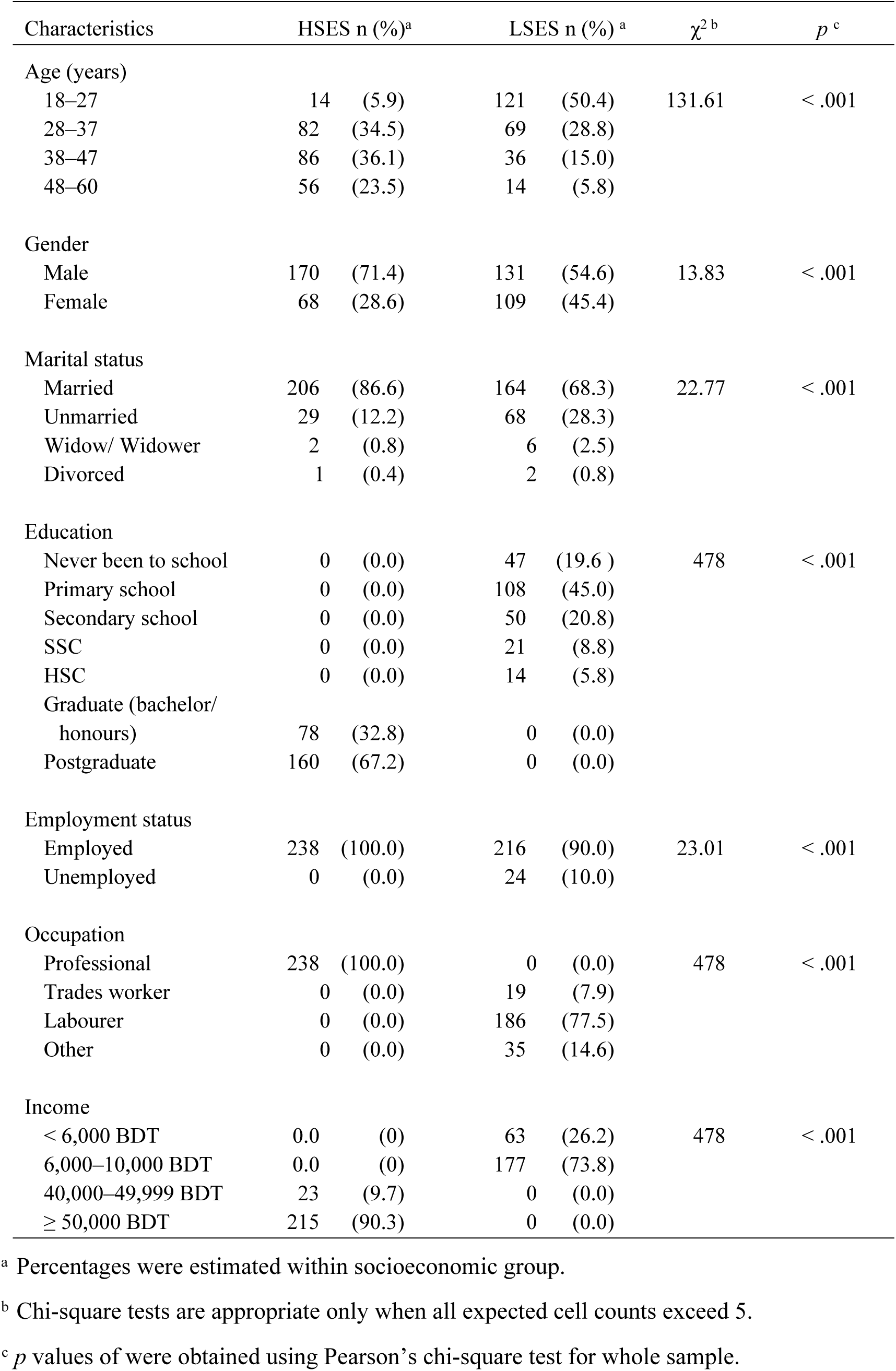
Demographic and occupation distribution for participants from each SES group (HSES and LSES)

These demographic characteristics provide important insights into the composition of the two SES groups in terms of occupation, age, and gender distribution. Understanding these demographic factors is crucial for interpreting the results and implications of the study about non-communicable diseases and population health.

### Knowledge about Coronary Heart Disease

Knowledge about Coronary Heart Disease (CHD) was assessed among participants from high socioeconomic status and low socioeconomic status groups. A significant difference was found between the knowledge scores of HSES (M = 18.77, SD = 4.0) and LSES (M = 9.66, SD = 4.1) participants, t(475.9) = 24.66, p < .001, d = 2.25. Despite this difference, none of the participants in either group were categorized as having a good or very good level of knowledge about CHD. Among HSES participants, 75.6% were classified as having a poor level of knowledge, and 60.0% of the LSES participants were categorized as having very poor knowledge.

Regarding specific knowledge aspects, approximately half of the participants from both socioeconomic groups were unaware that heart attack/heart disease is the leading cause of death in Bangladesh. However, the majority of participants from both groups (HSES 84.5%, LSES 74.6%) were aware that CHD is a long-term disease.

In the HSES group, 26.9% of participants identified at least three risk factors, and 23.5% identified at least four. In contrast, among LSES participants, 32.1% were unable to identify a single risk factor for CHD, and the same percentage mentioned only one risk factor. Notably, approximately 30% of participants from both groups mentioned factors not recognized by the World Health Organization (WHO) as CHD risk factors, including work stress, food adulteration, drug addiction, air pollution, and infection. These factors were not included in the total knowledge score. The results showed that HSES participants consistently demonstrated significantly better knowledge about CHD pathophysiology and risk factors compared to LSES participants. The only difference observed in symptoms of a heart attack was that HSES participants were more aware that sweating is a symptom.

Participants from both SES groups listed a maximum of five symptoms associated with a heart attack. In the HSES group, 26.1% knew at least one symptom, while in the LSES group, 24.2% knew at least one symptom. Additionally, 29.4% of HSES participants and 31.7% of LSES participants knew at least two symptoms. Furthermore, a significant proportion of participants believed that a person could be unaware of having CHD, with 98.3% of HSES participants and 75% of LSES participants expressing this belief.

Regarding prevention of CHD, there was a significant SES difference between those who believed that CHD could be prevented, with a higher percentage in the HSES group (92.9%) compared to the LSES group (62.9%), (p < .001). Similarly, there was a significant SES difference between those who thought CHD could be prevented by controlling or avoiding risk factors, with a higher percentage in the HSES group (88.2%) compared to the LSES group (35.8%), (p < .001).

Almost all participants from both groups believed that CHD was treatable, with 99.6% in the HSES group and 95% in the LSES group expressing this belief. When asked about treatment options, the majority of participants mentioned medication (HSES 67.1%, LSES 69.3%). However, there was a significant difference between the groups, with a higher percentage of HSES participants (63.7%) considering surgery as a treatment option compared to LSES participants (9.2%), (p < .001). Table 2 provides a comprehensive overview of participant responses to knowledge questions related to CHD pathophysiology, risk factors, and symptoms of a heart attack. HSES vs LSES on CHD knowledge domains were compared for multiple-response items using goodness-of-fit χ² with Bonferroni correction. These findings highlight the areas where targeted education and awareness campaigns can be implemented to improve knowledge about CHD among individuals from different socioeconomic backgrounds. These results highlight the disparities in knowledge levels between different socioeconomic groups and emphasize the need for targeted interventions to improve awareness and understanding of CHD, particularly among individuals with lower socioeconomic status.

**Table 2.** Participant knowledge of CHD pathophysiology, risk factors, and symptoms, including differences between HSES and LSES groups.

| Knowledge category | HSES: *n* (%)<sup>a</sup> | LSES: *n* (%)<sup>a</sup> | $\chi^2$ | *p*<sup>b</sup> |
| --- | --- | --- | --- | --- |
| CHD pathophysiology |  |  |  |  |
| Understanding of CHD |  |  |  |  |
| Pathological process | 148 (62.2) | 26 (10.8) | 86.67 | < .001 |
| Risk factors | 102 (42.9) | 57 (23.8) | 13.08 | < .001 |
| Do not know | 1 (0.4) | 27 (11.2) | 24.14 | < .001 |
| CHD sequelae |  |  |  |  |
| Malfunction of heart due to inappropriate blood circulation resulting from narrowing / blockage of arteries as a result of fat deposition | 195 (81.9) | 54 (13.0) | 160.66 | < .001 |
| Do not know | 23 (9.7) | 179 (74.6) | 119.24 | < .001 |
| The five most commonly mentioned CHD risk factors<sup>c</sup> |  |  |  |  |
| Unhealthy diet | 199 (83.6) | 71 (29.6) | 61.83 | < .001 |
| Mental stress | 135 (56.7) | 102 (42.5) | 4.88 | .027 |
| Smoking | 126 (52.9) | 48 (20.0) | 35.69 | < .001 |
| Physical inactivity | 128 (53.8) | 18 (7.5) | 83.78 | < .001 |
| High cholesterol | 110 (46.2) | 5 (2.1) | 96.60 | < .001 |
| The five most commonly mentioned symptoms of heart attack<sup>d</sup> |  |  |  |  |
| Chest pain | 216 (90.8) | 199 (82.9) | 0.70 | .404 |
| Breathlessness | 63 (26.5) | 67 (27.9) | 0.09 | .766 |
| Sweating | 88 (37.0) | 10 (4.2) | 62.72 | < .001 |
| Palpitation | 16 (6.7) | 31 (12.9) | 4.66 | .031 |
| Dizziness | 25 (10.5) | 19 (7.9) | 0.87 | .349 |

<sup>a</sup> Percentages are calculated within each socioeconomic status. *n* shown for each cell
<sup>b</sup> For multiple response questions, *p* values were estimated by comparing the percentages of HSES and LSES columns using the goodness-of-fit chi-square test with *df* = 1.
<sup>c</sup> Significance was set at .002 with a Bonferroni adjustment given the full complement of 21 CHD risk factors that participants mentioned.
<sup>d</sup> Significance was set at .003 with a Bonferroni adjustment, given the full complement of 16 heart attack symptoms that participants mentioned

### Awareness about CHD

Single-response awareness categories across SES were analysed using Pearson’s χ² with within-table denominators (Table 3). There was a significant difference between the awareness scores of HSES (M = 9.50, SD = 3.1) and LSES (M = 6.06, SD = 2.6) participants, t(476) = 13.22, p < .001, d = 1.20. Only 19.4% of the HSES participants had either a good or very good level of awareness about CHD, but this was noticeably higher than the 2.1% of LSES participants within those two levels of awareness. Almost half (45.8%) of the HSES participants were categorized as having a moderate level of awareness. In contrast, 56.7% of the LSES participants had a poor level of CHD awareness.

**Table 3.** Participant awareness of CHD-related conditions, diagnosis history, family history, and community exposure across SES groups.

| Category | HSES: *n* (%)<sup>a</sup> | LSE: *n* (%)<sup>a</sup> | $\chi^2$ | *p*<sup>b</sup> |
| --- | --- | --- | --- | --- |
| Awareness about having medical conditions related to CHD |  |  |  |  |
| Aware of having a medical condition | 66 (27.7) | 20 (8.3) | 39.50 | < .001 |
| Aware of not having a medical condition | 120 (50.4) | 182 (75.8) |  |  |
| Unaware of having or not having | 52 (21.8) | 38 (15.8) |  |  |
| Awareness about prediagnosed CHD |  |  |  |  |
| Aware of having prediagnosed CHD | 13 (5.5) | 12 (5.0) | 0.000 | .983 |
| Aware of not having prediagnosed CHD | 225 (94.5) | 228 (95.0) |  |  |
| Awareness about having a family history of CHD |  |  |  |  |
| Aware of having | 91 (38.2) | 44 (18.3) | 44.36 | < .001 |
| Aware of not having | 139 (58.4) | 149 (62.1) |  |  |
| Unaware either way | 8 (3.4) | 47 (19.6) |  |  |
| Awareness of someone with CHD in the community<sup>c</sup> |  |  |  |  |
| Aware of having | 187 (78.6) | 60 (25.0) | 139.15 | < .001 |
| Aware of not having | 23 (9.7) | 59 (24.6) |  |  |
| Unaware either way | 28 (11.8) | 121 (50.4) |  |  |

<sup>a</sup> Percentages were estimated within each socioeconomic status. *n* shown for each cell.
<sup>b</sup> *Pearson's $\chi^2$ for single-response items; goodness-of-fit $\chi^2$ for multiple-response items. Bonferroni thresholds applied for multiple-response domains (see caption). $\chi^2$ not performed where expected counts <5.*

Most HSES participants (61.8%) were aware of their own CHD risk, in contrast to only 25.4% of the LSES participants (p < .001). Participants in both HSES and LSES groups who were aware of having one or more risk factors stated that mental stress was the most common factor (32% and 56.7%, respectively). More than 65% of HSES participants, in contrast to approximately 25% of LSES participants, reported at least one personal risk factor they were aware of.

There was a significant SES difference between participants who indicated they took preventive action in relation to CHD (HSES 57.1% vs LSES 10.4%, p < .001), and among each of those groups, 72.3% of HSES and 60% of LSES participants stated that they were leading a healthy lifestyle as a preventive measure. Less than half of the participants from both groups (HSES 36.8% and LSES 40%) reported that they took preventive action in relation to their overall health, such as leading a healthy lifestyle, visiting doctors regularly and having regular health checkups, and taking medicine if required, and 14.7% of HSES participants stated that they took preventive action because they already had chronic diseases such as CHD, HTN, diabetes, high cholesterol, or stroke.

Participant responses associated with awareness about having medical conditions related to CHD, prediagnosed CHD, having a family history of CHD, and knowing a person with CHD in the community are presented in Table 3, where the most noticeable differences are that LSES participants were more likely to say they were aware of not having a CHD-related condition. Still, HSES participants were more likely to be aware that their family had a history of CHD and also to be aware of someone in the community who had CHD.

### Lifestyle Practices

Although the participant levels of knowledge and awareness were not high, approximately half of the HSES (57.1%) and LSES (50.4%) groups had lifestyle-practice scores that were categorized as good. Approximately one quarter of the participants in both HSES (28.6%) and LSES (27.9%) groups had lifestyle-practice scores rated as very good. There was no significant difference between lifestyle-practice scores between HSES (M = 3.63, SD = 0.55) and LSES (M = 3.53, SD = 0.68) participants, t(458.89) = 1.696, p = .090.

Participant responses relating to lifestyle practices are presented in Table 4. SES differences in behavioural practices are presented with Pearson’s χ² and standardized denominators for each item. The results showed that neither group had a high percentage of tobacco smokers; HSES participants tended to be ex-smokers, and LSES participants tended to be nonsmokers. LSES participants were more likely to be among those who chewed tobacco. A much higher percentage of LSES participants were physically active at work but undertook no exercise when not at work. Both groups exhibited a similar pattern, with almost all participants consuming fewer than five servings of fruits and vegetables daily. In separate analyses, a significant difference was seen in smoking with respect to gender, where none of the female participants from either group were current smokers or ex-smokers. However, a greater percentage of the females chewed tobacco compared to males. Among the female participants, a higher percentage was from the LSES group. Alcohol consumption was very low in both groups. Fast-food consumption was significantly higher among HSES participants, while extra salt intake in meals was significantly higher among LSES participants. A notable proportion of LSES participants did not consume fish, meat, or protein alternatives daily and consumed extra salt with their meals.

**Table 4.** Lifestyle behaviors of participants, including tobacco use, physical activity, and dietary habits across SES groups.

| Characteristics | HSES: *n* (%)<sup>a</sup> | LSES: *n* (%)<sup>a</sup> | $\chi^2$ | *p*<sup>b</sup> |
| --- | --- | --- | --- | --- |
| Tobacco smoking and chewing |  |  |  |  |
| Smoking |  |  |  |  |
| Smoker | 45 (18.9) | 47 (19.6) | 12.00 | .002 |
| Nonsmoker | 167 (70.2) | 186 (77.5) |  |  |
| Ex-smoker | 26 (10.9) | 7 (2.9) |  |  |
| Chewing |  |  |  |  |
| Tobacco chewer | 14 (5.9) | 44 (18.3) | 19.63 | < .001 |
| Tobacco nonchewer | 222 (94.1) | 192 (81.7) |  |  |
| Tobacco ex-chewer | 1 (0.4) | 4 (1.7) |  |  |
| Physical activity of participants |  |  |  |  |
| Physically active at work |  |  |  |  |
| Yes | 19 (8.0) | 117 (48.8) | 95.57 | < .001 |
| No | 219 (92.0) | 123 (51.2) |  |  |
| Level of physical activity apart from work |  |  |  |  |
| Sedentary | 70 (29.4) | 111 (46.2) | 27.60 | < .001 |
| Light | 54 (22.7) | 51 (21.2) |  |  |
| Moderate | 76 (31.9) | 69 (28.8) |  |  |
| Strenuous | 38 (16.0) | 9 (3.8) |  |  |
| Fruit and vegetables |  |  |  |  |
| < 5 serves of fruits and vegetables each day | 220 (92.4) | 232 (96.7) | 0.32 | .572 |

<sup>a</sup> Percentages apply within each socioeconomic status. *n* shown for each cell.
<sup>b</sup> *Pearson's $\chi^2$ for single-response items; goodness-of-fit $\chi^2$ for multiple-response items. Bonferroni thresholds applied for multiple-response domains (see caption). $\chi^2$ not performed where expected counts < 5.*

### Correlations between knowledge, awareness, and lifestyle practices

Knowledge and awareness about CHD were strongly correlated with each other when the total sample was considered (rs = .601, p < .001), indicating a positive association between the two variables. However, when the socioeconomic status (SES) groups were considered separately, there were only moderate associations between knowledge and awareness. Specifically, in the HSES participants, the correlation between knowledge and awareness was rs = .38, p < .001, while in the LSES participants, the correlation was rs = .45, p < .001. These results suggest that the relationship between knowledge and awareness varied depending on the socioeconomic status of the participants.

Interestingly, the lifestyle practice score was not significantly correlated with CHD knowledge or CHD awareness in either SES group. This indicates that there was no significant association between participants’ level of knowledge or awareness about CHD and their lifestyle practices related to the disease. In other words, participants’ knowledge and awareness levels did not directly translate into changes in their lifestyle practices.

These findings suggest that while there is a strong positive association between knowledge and awareness about CHD, the impact of this knowledge and awareness on participants’ actual lifestyle practices may be influenced by other factors beyond their level of understanding. Further investigation is needed to identify the factors that contribute to participants’ lifestyle practices and explore the complex relationship between knowledge, awareness, and behavior change in the context of CHD.

### Multiple linear regression analyses

Standard (simultaneous) multiple linear regression analyses were used to determine whether SES, age, and gender could predict knowledge and awareness, and also whether SES, knowledge, awareness, age, and gender could predict lifestyle practices. Although SES was the primary research focus, age and gender were regarded as appropriate inclusions because of the earlier results showing significant differences in age between the two socioeconomic strata, as well as gender differences in the HSES participants.

### Predicting knowledge

In predicting participant knowledge, four models attained statistical significance according to their *F* values, and an AIC Δ < 2 relative to each other. Among the models, Model 2 (Table 5) was assessed as being the most satisfactory because within it both SES and age were statistically significant predictors of knowledge, *t* = 20.71, *p* < .001 and *t* = 2.14, *p* < .033, respectively, and, for that model, the adjusted *R^2^*value indicated that 56.33% of the variability in knowledge about CHD could be explained by SES and age. As can be seen from entries in Table 5, the standardized β coefficients indicate that SES was a considerably stronger predictor of knowledge than was age (|0.72| versus |0.07|, respectively). From the unstandardized beta coefficients in this model (−8.68 and +0.04 for SES and age, respectively), the higher a participant’s knowledge about CHD, the more likely that participant was to come from the HSES and to be older.

**Table 5.** Regression Summary Table for Knowledge, Awareness, and Lifestyle Practices Based on Results from the Most Satisfactory Models.

| Model | Unstandardized coefficients |  | Standardized coefficients |  |  |
| --- | --- | --- | --- | --- | --- |
| | *B* | Standard error | $\beta$ | *t* | *p* |
| Knowledge<br>(Model 2) |  |  |  |  |  |
| Constant | 17.16 | 0.80 | - | 21.54 | < .001 |
| SES<sup>a</sup> | -8.68 | 0.42 | -0.72 | 20.71 | < .001 |
| Age | 0.04 | 0.19 | 0.07 | 2.14 | .033 |
| Awareness<br>(Model 1) |  |  |  |  |  |
| Constant | 8.67 | 0.26 | - | 32.84 | < .001 |
| SES<sup>a</sup> | -3.24 | 0.26 | -0.49 | 12.50 | < .001 |
| Gender<sup>b</sup> | 1.16 | 0.27 | 0.17 | 4.32 | < .001 |
| Lifestyle practices<br>(Model 1) |  |  |  |  |  |
| Constant | 4.45 | 0.11 | - | 38.43 | < .001 |
| SES<sup>a</sup> | -0.32 | 0.06 | -0.03 | 5.17 | < .001 |
| Age | -0.02 | 0.00 | -0.37 | 7.51 | < .001 |

<sup>a</sup> Negative coefficients are associated with LSES participants.
<sup>b</sup> Positive coefficients are associated with males.

### Predicting awareness

In predicting participant awareness, four models again attained statistical significance according to their *F* values, but only the first two had an AIC Δ < 2 relative to each other. Models 3 and 4 had an AIC Δ > 10 relative to Model 2, so only the first and second models were considered further. Within Model 1, SES and gender were both significant predictors of awareness, *t* = 12.50, *p* < .001, and *t* = 4.32, *p* < .001, respectively. Model 2 contained the additional variable of age, but it was not significant, *t* = 0.15, *p* = .882. Therefore, Model 1 was regarded as the most satisfactory. The adjusted *R^2^*value in this model indicated that SES and gender could explain 29.30% of the variability in awareness. Furthermore, the standardized β coefficients of |0.49| and |0.17| indicate that SES was a stronger predictor of awareness than was gender (Table 5). From the unstandardized beta coefficients in this model (−3.24 and +1.16 for SES and gender, respectively), the higher a participant’s awareness about CHD, the more likely that participant came from the HSES and was male.

### Predicting lifestyle practices

Analyses for determining the extent to which participant lifestyle practices could be predicted by their SES, knowledge, awareness, age, and gender initially generated 28 models, but only the four models that provided the strongest predictive outcomes were considered for subsequent analyses. All these four models attained statistical significance according to their *F* values, and all had an AIC Δ < 2 relative to each other, so they were indistinguishable in predictive power. Based on AICs, the most effective means of predicting lifestyle practices were SES and age in Model 1. In that model, both variables were statistically significant predictors of lifestyle practices, *t* = 5.17, *p* < .001, and *t* = 7.51, *p* < .001, respectively. The adjusted *R^2^*value indicated that SES and age could explain 10.78% of the variability in the participant lifestyle practices. As can be seen from the standardized β coefficients of |0.37| and |0.03| in Table 5, age carried greater predictive power than did SES. In Model 1, the unstandardized beta coefficients of −0.32 and −0.02 for SES and age, respectively, indicate that better lifestyle practices are more likely when a participant is from the HSES group and younger.

## Discussion

A study conducted in Bangladesh revealed a lack of knowledge about coronary heart disease (CHD), especially among individuals from lower socioeconomic status (LSES) [32]. In contrast to a study in Malaysia, where 70% had good knowledge, no participants in Bangladesh reached that level. Limited health education programs on noncommunicable diseases (NCDs) and restricted access to information and education likely contributed to poorer CHD knowledge among LSES individuals [33]. Participants had a better understanding of CHD prevention than risk factors and symptoms. Still, there was a lack of awareness regarding heart attack symptoms beyond chest pain, suggesting the need for improved education [11, 32, 33, 34]. LSES participants had less awareness of surgical interventions due to limited healthcare access [11, 32]. Targeted health education programs are crucial to improve CHD knowledge, especially among individuals from lower socioeconomic backgrounds, addressing education gaps and healthcare access [32, 33, 34, 35, 36]. Both socioeconomic groups lacked knowledge about heart attack symptoms beyond chest pain, highlighting the need for increased awareness. The study’s findings underscore the importance of focused health education initiatives to enhance CHD knowledge in the general population, with a specific focus on individuals from lower socioeconomic backgrounds. Efforts should concentrate on increasing awareness of CHD risk factors, symptoms, and prevention while addressing educational disparities and improving healthcare access for those with lower socioeconomic status.

Awareness levels about coronary heart disease (CHD) vary across different populations. Studies in Saudi Arabia, the United States, and Jamaica revealed significant differences in awareness percentages [12, 16, 17]. In the present study, both high and low socioeconomic status (HSES and LSES) participants lacked CHD awareness. LSES individuals had significantly lower awareness of their own CHD risk factors compared to HSES participants. Only 10.4% of LSES participants were aware of the importance of preventive actions, contrasting with a study in the USA where nearly 90% reported annual checkups. Limited education, knowledge, and access to healthcare likely contributed to the lower engagement in preventive actions among LSES participants. Their awareness of CHD relied on external sources after experiencing symptoms and visiting primary healthcare facilities. LSES participants also demonstrated lower awareness of pre-diagnosed conditions like hypertension, diabetes, and high cholesterol, attributed to factors such as age, education, diagnosis rates, and limited access to healthcare providers. Similar findings were reported in studies conducted in Bangladesh and India [32]. Both socioeconomic groups expressed intentions to seek medical help in the event of a heart attack, aligning with findings from Nepal [25].

The majority of participants from high socioeconomic status (HSES) and low socioeconomic status (LSES) groups demonstrated good lifestyle practices, with 87.3% overall. HSES participants scored higher in specific domains like ex-smoking, while LSES participants scored higher in tobacco chewing [32]. Smoking prevalence was similar to an earlier survey in Bangladesh, with higher prevalence among LSES individuals. Tobacco chewing was more common among LSES participants, especially women [40]. Alcohol consumption was minimal in both groups [41]. In terms of dietary habits, both groups consumed fewer than five servings of fruits and vegetables daily, with LSES participants having a higher percentage of extra salt intake [8].

Knowledge and awareness of CHD were strongly correlated in both SES groups, indicating that individuals with better CHD knowledge also had higher awareness. Similar findings have been reported for HTN and diabetes in India [43]. However, there were no significant associations between CHD knowledge or awareness and lifestyle practices within either SES group. However, other factors, including environmental factors, such as concerns about food adulteration, as well as socioeconomic and sociocultural factors, were also evident, with LSES participants being more physically active at work and less likely to consume fast food, while HSES participants had more sedentary occupations and were more inclined to consume fast food. Alcohol consumption was very low due to cultural and religious restrictions.

Socioeconomic status (SES), gender, and age were significant predictors of knowledge, awareness, and lifestyle practices related to CHD. SES, particularly educational attainment, played a crucial role in predicting knowledge about CHD, consistent with findings from Canada and Jordan [45, 46]. Men had higher awareness levels, possibly due to higher education levels in the LSES group and cultural influences [47, 48, 49]. However, neither knowledge nor awareness significantly predicted lifestyle practices, in line with mixed findings in previous research [59]. Instead, SES and age emerged as significant predictors, with LSES participants and older individuals exhibiting poorer lifestyle practices [34, 51]. This study contributes to understanding the associations between knowledge, awareness, and lifestyle practices in CHD, emphasizing socioeconomic factors, gender, and age as important predictors. Limitations include gender disparity in participant recruitment, partial validation of scoring procedures, and opportunities for refining the conception, measurement, and analysis of lifestyle practices [52, 53].

This study has several limitations that should be considered when interpreting the findings. Recruitment was limited to Dhaka and employed various sampling frames, including office workplaces for HSES and selected slum areas for LSES, as well as proximity-based selection. This approach made the findings more applicable to urban Bangladesh, although selection bias cannot be ruled out. The two SES groups also differed in age and gender (HSES participants were older and more often male). Although the statistical analyses adjusted for these factors, some residual confounding may still exist. Outcomes were assessed in single, face-to-face interviews using a questionnaire that, despite careful development, translation, local review, and pilot testing, remains susceptible to recall bias, social desirability bias, and misclassification of knowledge, awareness, and practices. Health indicators combined self-report (for diagnosed conditions) with point measurements (blood pressure and random glucose), which may misclassify chronic conditions when based on a single reading rather than clinical confirmation. Multiple-response knowledge items also required Bonferroni correction. This is an appropriate statistical process for controlling multiplicity but may increase the risk of Type II error and hide smaller between-group differences. Finally, the cross-sectional, single-timepoint design prevents causal inference, especially with respect to the lack of association between knowledge/awareness and lifestyle, which indicates that unmeasured structural or contextual factors may influence behavior despite higher knowledge levels.

## Conclusion

In this urban Bangladeshi sample, people of higher socioeconomic status showed greater knowledge and awareness of coronary heart disease (CHD) than those of lower status. However, these qualities were not linked to healthier lifestyle habits, indicating that information alone is not enough to change behavior, and that interventions should combine health education, especially tailored to lower-income groups and include structural supports that reduce barriers to change such as accessible screening and counselling, tobacco control and cessation support, healthier food environments, and pathways for managing hypertension and diabetes. Due to the cross-sectional, Dhaka-based design, causal inference and national generalizability are limited. However, the findings highlight actionable priorities for urban CHD prevention.

## Data Availability

All data produced in the present study are available upon reasonable request to the authors

## References

1. World Health Organization. Global Health Estimates 2019: Deaths by Cause, Age, Sex, by Country and by Region, 2000-2019. World Health Organization; 2020. Accessed May 10, 2023. https://www.who.int/data/gho/data/themes/mortality-and-global-health-estimates/ghe-leading-causes-of-death

2. Ibrahim MM, Damasceno A. Hypertension in developing countries. Lancet. 2012;380(9841):611–619. doi:10.1016/S0140-6736(12)60861-7

3. World Health Organization. Noncommunicable diseases country profiles 2018. World Health Organization; 2018. Accessed May 10, 2023. https://www.who.int/nmh/publications/ncd-profiles-2018/en/

4. Rahman MS, Akter S, Abe SK, et al. Awareness, treatment, and control of hypertension among the adult population in Bangladesh. Hypertens Res. 2019;42(10):1666–1678. doi:10.1038/s41440-019-0274-2

5. Puska P, Vartiainen E, Tuomilehto J, et al. Changes in premature deaths in Finland: successful long-term prevention of cardiovascular diseases. Bull World Health Organ. 1998;76(4):419–425.

6. Biswas T, Islam A, Rawal LB, et al. Socioeconomic inequality in the prevalence of noncommunicable diseases in low– and middle-income countries: Results from the World Health Survey. BMC Public Health. 2016;16:1–9. doi:10.1186/s12889-016-3734-3

7. World Health Organization. Global status report on alcohol and health 2018. World Health Organization; 2018. Accessed May 10, 2023. https://www.who.int/publications/i/item/9789241565639

8. World Health Organization. Global status report on noncommunicable diseases 2014. World Health Organization; 2014. Accessed May 10, 2023. https://www.who.int/nmh/publications/ncd-status-report-2014/en/

9. World Health Organization. Noncommunicable diseases progress monitor 2017. World Health Organization; 2017. Accessed May 10, 2023. https://www.who.int/nmh/publications/ncd-progress-monitor-2017/en/

10. Shrivastava SR, Shrivastava PS, Ramasamy J. Role of self-care in management of coronary heart disease. J Cardiovasc Dis Res. 2011;2(4):189–192. doi:10.4103/0975-3583.89802

11. Redman BK. The Practice of Patient Education: A Case Study Approach. Mosby; 2006.

12. Becker MH. The health belief model and personal health behavior. Health Educ Monogr. 1974;2(4):324–473. doi:10.1177/109019817400200405

13. Stringhini S, Sabia S, Shipley M, et al. Association of socioeconomic position with health behaviors and mortality. JAMA. 2010;303(12):1159–1166. doi:10.1001/jama.2010.297

14. Chow CK, Lock K, Teo K, et al. Environmental and societal influences acting on cardiovascular risk factors and disease at a population

15. Biswas T, Islam A, Pervin S, et al. Socio-economic inequality of chronic noncommunicable diseases in Bangladesh. PLoS One. 2016;11(11):e0167140. doi:10.1371/journal.pone.0167140

16. Bleich SN, Koehlmoos TLP, Rashid M, et al. Non-communicable disease prevention in Bangladesh: a policy analysis. Lancet. 2013;382:S12. doi:10.1016/S0140-6736(13)62359-8

17. Chowdhury MZI, Anik AM, Farhana Z, et al. Prevalence of risk factors for non-communicable diseases in Bangladesh: results from STEPS survey 2010. Indian J Public Health. 2016;60(1):17–25. doi:10.4103/0019-557X.177281

18. Khan JR, Biswas T. Rising rural body-mass index is the main driver of the global obesity epidemic in adults. Nature. 2017;549(7672):260-264. doi:10.1038/nature23682

19. Jahan Y, Nahar S, Hossain S, et al. Tobacco use in Bangladesh: prevalence, trends, and determinants. J Health Popul Nutr. 2019;38(1):1–15. doi:10.1186/s41043-019-0173-2

20. Zaman MM, Bhuiyan MR, Karim MN, et al. Clustering of non-communicable diseases risk factors in Bangladeshi adults: an analysis of STEPS survey 2013. BMC Public Health. 2015;15:659. doi:10.1186/s12889-015-2038-2

21. WHO. 2018 National STEPS Survey for Non-communicable Diseases Risk Factors in Bangladesh. Dhaka: WHO; 2022. Dhaka, Bangladesh: National Institute of Preventive and Social Medicine (NIPSOM) Mohakhali, Dhaka 1212 2022.

22. Riaz BK, Islam MZ, Islam A, Zaman MM, Hossain MA, Rahman MM, et al. Risk factors for non-communicable diseases in Bangladesh: findings of the population-based cross-sectional national survey 2018. BMJ Open. 2020;10(11):e041334.

23. Islam AK, Majumder AA. Coronary artery disease in Bangladesh: a review. Indian Heart J. 2013;65(4):424–35.

24. Siddique AB, Hosen MS, Akter H, Hossain SM, Al Mamun M. Assessment of knowledge, attitudes, and practices regarding cardiovascular diseases (CVDs) among older individuals of rural Bangladesh: findings from a face-to-face interview. Frontiers in public health. 2024;12:1336531.

25. Rahman DMM, Akter S, Zohora F, Shibly D. Public knowledge of cardiovascular disease and its risk factors in Tangail, Bangladesh: a cross-sectional survey. International Journal of Community Medicine and Public Health. 2019;6:1838.

26. Mistry SK, Hossain MB, Parvez M, Gupta RD, Arora A. Prevalence and determinants of hypertension among urban slum dwellers in Bangladesh. BMC Public Health. 2022;22(1):2063.

27. Khanam F, Hossain MB, Mistry SK, Afsana K, Rahman M. Prevalence and Risk Factors of Cardiovascular Diseases among Bangladeshi Adults: Findings from a Cross-sectional Study. J Epidemiol Glob Health. 2019;9(3):176–84.

28. Chowdhury MZI, Rahman M, Akter T, Akhter T, Ahmed A, Shovon MA, et al. Hypertension prevalence and its trend in Bangladesh: evidence from a systematic review and meta-analysis. Clin Hypertens. 2020;26:10.

29. Mirza A-S, Aslam S, Perrin K, Curtis T, Stenback J, Gipson J, et al. Knowledge, attitudes and practices among patients with coronary artery disease in Dhaka, Bangladesh. Int J Community Med Public Health. 2016;3(10):2740–8.

30. Saeed O, Gupta A, Dhawan N, et al. Knowledge, awareness, and perception of myocardial infarction and its treatment among urban and rural populations in six low– and middle-income countries: A cross-sectional study. PLoS One. 2018;13(6):e0196676. doi:10.1371/journal.pone.0196676.

31. Dudala S. Updated Kuppuswamy’s socioeconomic scale for 2012. J Dr NTR Univ Health Sci. 2013;2(3):201–2. 10.4103/2277-8632.117195

32. Muhamad R, Yahya R,Yusoff, HM. Knowledge, attitude and practice on cardiovascular disease among women in north-east coast Malaysia. Int J Collab Res Intern Med Public Health. 2012;4(1):85–98.

33. Jafary FH, Aslam F, Mahmud H, Waheed A, Shakir M, Afzal A, et al. Cardiovascular health knowledge and behavior in patient attendants at four tertiary care hospitals in Pakistan – a cause for concern. BMC Public Health. 2005;5:124. 10.1186/1471-2458-5-124

34. Vaidya A, Aryal UR, Krettek A. Cardiovascular health knowledge, attitude and practice/behaviour in an urbanising community of Nepal: a population-based cross-sectional study from Jhaukhel-Duwakot Health Demographic Surveillance Site. BMJ Open. 2013;3(10):e002976. 10.1136/bmjopen-2013-002976

35. Gurjar BR, Butler TM, Lawrence MG, Lelieveld J. Evaluation of emissions and air quality in megacities. Atmos Environ. 2008;42(7):1593–606. 10.1016/j.atmosenv.2007.10.048

36. Mahmood S, Ali S. Shifting from infectious diseases to non-communicable diseases: a double burden of diseases in Bangladesh. J Public Health Epidemiol. 2013;5(11): 424–34.

37. Quasem IS, Shetye M, Alex SC, Nag AK, Sarma PS, Thankappan K R, Vasan RS. (2001). Prevalence, awareness, treatment and control of hypertension among the elderly in Bangladesh and India: A multicentre study. Bulletin of the World Health Organization, 79(6), 490–500.

38. Sultana P, Akter S, Rahman M, Alam S. Prevalence and predictors of current tobacco smoking in Bangladesh. Journal of Biometrics and its Application. 2015;1.

39. Islam FMA, Walton A. Tobacco smoking and use of smokeless tobacco and their association with psychological distress and other factors in a rural district in Bangladesh: a cross-sectional study. J Environ Public Health. 2019;2019:1424592. 10.1155/2019/1424592

40. Gupta PC, Ray CS. Smokeless tobacco and health in India and South Asia. Respirology. 2003;8(4):419–31. 10.1046/j.1440-1843.2003.00507.x

41. Islam JY, Zaman MM, Bhuiyan MR, Hasan MM, Ahsan HN, Rahman MM, et al. Alcohol consumption among adults in Bangladesh: Results from STEPS 2010. WHO South East Asia J Public Health. 2017;6(1):67–74. 10.4103/2224-3151.206168

42. Harun MA, Ahmed F, Maniruzzaman. Customer hospitality: the case of fast food industry in Bangladesh. World Journal of Social Sciences. 2013;3(6):88–104.

43. Chavan GM, Waghachavare VB, Gore AD, Chavan VM, Dhobale RV, Dhumale GB. Knowledge about diabetes and relationship between compliance to the management among the diabetic patients from rural area of Sangli District, Maharashtra, India. J Family Med Prim Care. 2015;4(3):439–43. 10.4103/2249-4863.161349

44. Yahya R, Muhamad R, Yusoff H. Association between knowledge, attitude and practice on cardiovascular disease among women in Kelantan, Malaysia. International Journal of Collaborative Research on Internal Medicine and Public Health. 2012;4:1507–23.

45. Kang Y, Yang IS, Kim N. Correlates of health behaviors in patients with coronary artery disease. Asian Nurs Res. 2010;4(1):45–55. 10.1016/S1976-1317(10)60005-9

46. Mukattash TL, Shara M, Jarab AS, Al-Azzam SI, Almaaytah A, Al Hamarneh YN. Public knowledge and awareness of cardiovascular disease and its risk factors: a cross-sectional study of 1000 Jordanians. Int J Pharm Pract. 2012;20(6):367–76. 10.1111/j.2042-7174.2012.00208.x

47. Ahmed SM, Adams AM, Chowdhury M, Bhuiya A. Gender, socioeconomic development and health-seeking behaviour in Bangladesh. Soc Sci Med. 2000;51(3):361–71. 10.1016/S0277-9536(99)00461-X

48. Rumi MH, Makhdum N, Rashid MH, Muyeed A. Gender differences in service quality of Upazila Health Complex in Bangladesh. J Patient Exp. 2021;8:23743735211008304. 10.1177/23743735211008304

49. Walton LM, Schbley B. Gender equity theory and maternal mortality and morbidity in Bangladesh: a review article. J Health Ethics. 2012;9(1):8. 10.18785/ojhe.0901.08

50. Asril NM, Tabuchi K, Tsunematsu M, Kobayashi T, Kakehashi M. Predicting healthy lifestyle behaviours among patients with type 2 diabetes in rural Bali, Indonesia. Clin Med Insights Endocrinol Diabetes. 2020;13:1179551420915856. 10.1177/1179551420915856

51. Ammouri AA, Abu Raddaha AH, Tailakh A, Kamanyire J, Achora S, Isac C. Risk knowledge and awareness of coronary heart disease, and health promotion behaviors among adults in Oman. Res Theory Nurs Pract. 2018;32(1):46–62. 10.1891/1541-6577.32.1.46

52. Kazim MN, AbouMoussa TH, Al-Hammadi FA, Ali AA, Abedini FM, Ahmad FSM, et al. Population awareness of cardiovascular disease risk factors and health care seeking behavior in the UAE. Am J Prev Cardiol. 2021;8:100255. 10.1016/j.ajpc.2021.100255

53. Moniruzzaman M, Mostafa Zaman M, Islalm MS, Ahasan HA, Kabir H, Yasmin R. Physical activity levels in Bangladeshi adults: results from STEPS survey 2010. Public Health. 2016;137:131–8. 10.1016/j.puhe.2016.02.028

